# Utility and impact of inpatient pediatric physiologic monitoring

**DOI:** 10.1101/2020.07.26.20162438

**Authors:** Anand Gourishankar, Monaliza Evangelista, Misti Ellsworth, Jean Hsu

## Abstract

**Objective:** To study the practice and effect of monitoring pediatric patients on the hospital length of stay (LOS).

**Methods:** We conducted a cross-sectional observational study of pediatric patients in the general ward from October to December 2016. We recorded the use of cardiac, pulse-oximeter, or both, and physician order at the time of admission. We studied the proportions of monitoring on different patient groups. The median length of stay assessed for various modes of monitoring. We did regression analysis for the effect of cardiopulmonary monitoring, orders, and medical complexity on hospital length of stay.

**Results:** Among 398 patients, patients with cardiac monitor and pulse oximeter with orders were 68 % and 82%, respectively. The pulmonary group of patients had more monitoring than the neurology group of patients. LOS was shorter in patients without monitors; the median difference for the cardiac monitor was 1 day, and pulse oximeter was 0.5 days. Cardiac monitor order increased LOS by 22% (95% CI, 0.5% to 48%) and complex past medical history increased it by 25% (95% CI, 4% to 51%).

**Conclusion:** Our study highlights the variable practice in using monitors, demanding a standardized approach. The judicious use of monitoring reduces prolonged hospital stay.

## Introduction

The judicious use of monitoring patients in the hospital poses a challenge. Pediatric patients admitted to the hospital are ill most of the time. Some are either psychosocial or elective admissions. On these ill and well-appearing patients, the decision and practice to use continuous electrocardiography, respiratory, and pulse oximetry monitoring varies widely among the providers. The health care providers’ understanding, and behavior determine the use of monitors. While there is a clear benefit of monitoring in specific populations[1], generalizing all inpatient use can cause harm, directly or indirectly (for example, alarm fatigue, unnecessary noise, false alarms, desensitization, nonactionable).[2] Unnecessary monitoring can be costly.

Direct bedside assessment at predetermined intervals is ideal, but a balance between continuous monitoring and physical evaluation is imperative. The assessment varies among specialties and locations within the hospital. There is an irregularity in either starting or discontinuing the monitors, and many factors influence their decisions.[3] There are gaps in health care providers’ knowledge and nurses’ expectation for placing monitors needs improvement. The aim was to study the practice and the effect of monitoring pediatric patients on the hospital length of stay.

## Methods

Patients admitted, between October and December 2016, to a single tertiary care center were enrolled in the cross-sectional study. The pediatric ward has three-floor levels with two are general ward, and one is intermediate acute care. The nursing ratio in the general ward can be 1:4-6 patients and 1:2-3 patients in the other ward. Institutional Review Board approved this study. A pediatric team consisting of attending and residents cared for the patients and comanaged with subspecialists. At the time of the study, community general pediatricians cross covered the hospitalist. Pediatric residents placed the admission orders. Guidelines for cardiac monitoring or pulse oximeter were not standardized and non-existent at the time of this study. The study authors collected the admission list every morning. These authors confirmed the application of pulse oximeter, cardiac monitor, or both at the bedside. Then, they cross-checked if the physician’s (resident or attending) ordered these monitoring. Patient data (recorded in Microsoft excel sheet, 2016) included location, age, date and time of admission/discharge, admission diagnosis, past medical history, presence of pulse oximeter or cardiac monitor, and documentation of physician order for monitoring. Data was stored in an encrypted University drive. Pediatric patients (ages up to 18yrs) admitted to the hospital medicine service were included. The exclusion criteria included patients admitted to the intensive care unit, surgical specialties that are not comanaged, and subspecialist care.

### Statistical analyses

Descriptive statistics were used for continuous variables such as age and length of stay. Proportions were reported in percentages. Interrater reliability (IRR) was analyzed using Fleiss’s Kappa.[4] The null hypothesis for IRR was kappa = 0, and kappa for “complex” and “simple” past medical history compared. A Chi-square test was applied to categorical data. Spearman correlation (rho) was applied to study the relationship between monitoring and the physician’s order. We analyzed non-parametric data with the Wilcoxon Mann Whitney test. Bivariate analysis was conducted on potential variables. Multiple linear regression was applied after verifying multicollinearity (variation inflation factor). Stepwise backward (hierarchical) log-linear regression method was used until a significant variable inferred for the length of hospital stay (primary outcome). Log transformation was applied to the skewed “length of stay’ variable. The final model was tested for autocorrelation (Durbin Watson Test), linearity, homodescasticity, constant variation, and outliers. There was one outlier but was found to be influential, and hence we included it in the final model. We compared the final model with the initial model using ANOVA. All calculations were statistically significant if p <0.05. Statistical computing was done with R statistical software (R-3.4.2).

## Results

The number of patients included in this study was 398. Forty-one were excluded due to incomplete data or missing data. Patients were admitted from our emergency department or accepted from outside health care facilities. The mean age was 5.2 years (SD, 5.6). The ages ranged from 3 days to 17 years. The ages by categories were infant 137 (30%), toddler 72 (20%), early childhood 51 (10%), middle childhood 61 (20%), and early adolescence 77 (20%); p = 0.03. Table 1 describes the number of patients by primary admission diagnosis of each (organ) system.

**Table 1.**
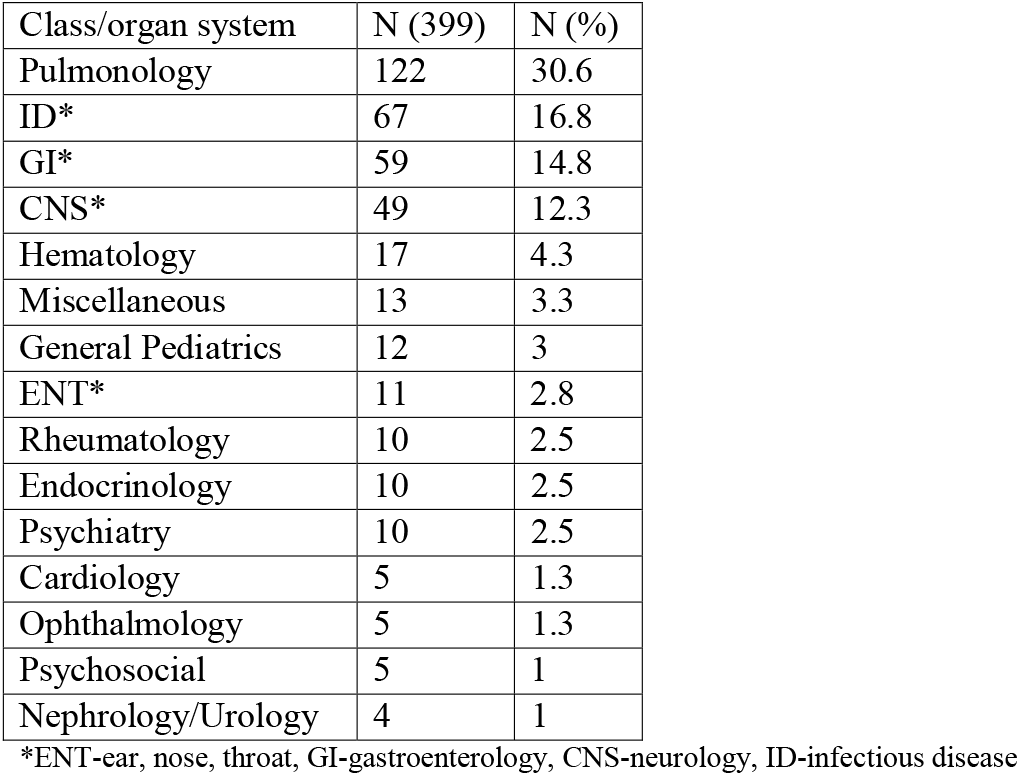
Admitted patients grouped by major systems

| Class/organ system | N (399) | N (%) |
| --- | --- | --- |
| Pulmonology | 122 | 30.6 |
| ID* | 67 | 16.8 |
| GI* | 59 | 14.8 |
| CNS* | 49 | 12.3 |
| Hematology | 17 | 4.3 |
| Miscellaneous | 13 | 3.3 |
| General Pediatrics | 12 | 3 |
| ENT* | 11 | 2.8 |
| Rheumatology | 10 | 2.5 |
| Endocrinology | 10 | 2.5 |
| Psychiatry | 10 | 2.5 |
| Cardiology | 5 | 1.3 |
| Ophthalmology | 5 | 1.3 |
| Psychosocial | 5 | 1 |
| Nephrology/Urology | 4 | 1 |

\*ENT-ear, nose, throat, GI-gastroenterology, CNS-neurology, ID-infectious disease

All authors classified the admission diagnoses into major systems, and any discrepancy was revisited until the final decision. Three authors (EM, HJ, EM) independently organized 398 patients into simple or complex medical conditions based on each patient’s past medical history. The interrater reliability, Fleiss’s kappa, was 0.51.

The proportion of patients with cardiac monitor and pulse oximeter with orders were 68% and 82%, respectively (Figure 1A). Two hundred and twenty patient patients (55%) had both types of monitors, and 148 (37%) did not have either of the two monitors. Medical complexities, as determined by the authors in “past medical history” and at “admission,” were 38% and 61%, respectively. There was significant association between the following physician orders; complex past medical history & cardiac monitor (p = 0.005), admission complexity & cardiac monitor (p < 0.001), admission complexity & pulse oximeter (p <0.001), and except complex past medical history & pulse oximeter (p = 0.29). The median age of cardiac monitor use with and without orders were 2 years and 1.5 years. The median age of pulse oximeter use was 1.5 years and 4 years, respectively (Figure 2).

**Figure 1.**
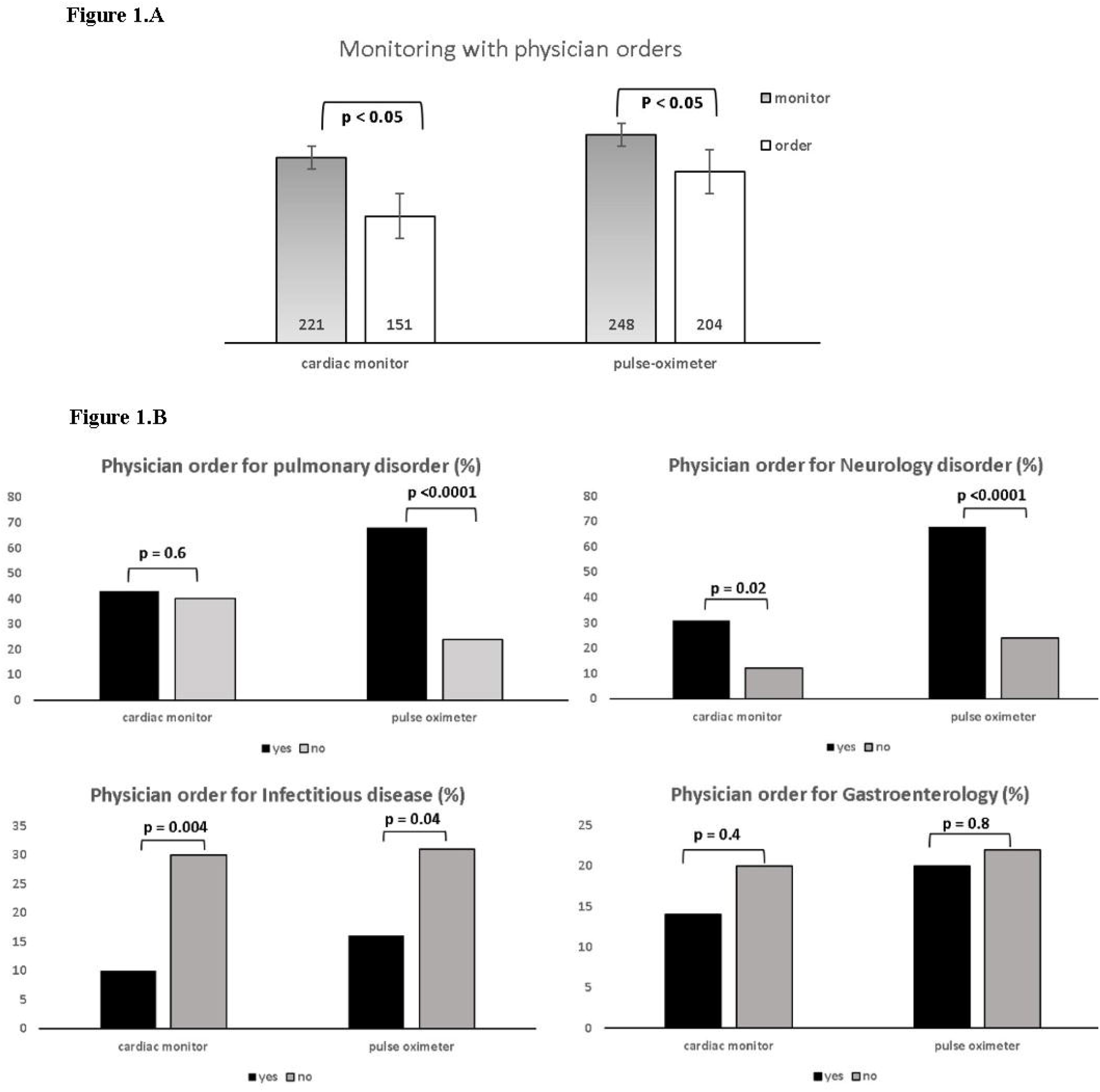
A.B. Cardiac and pulse oximeter monitor use based on the physician’s order

**Figure 2.**
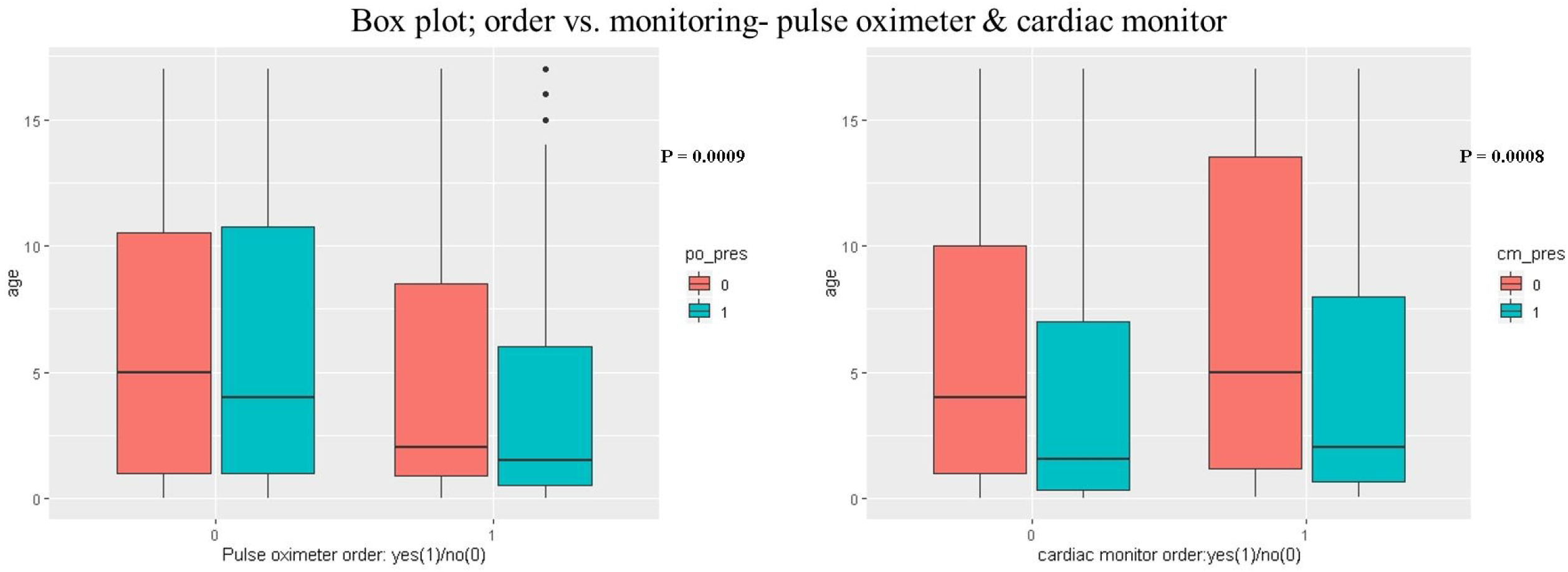
The median age among pulse oximeter and cardiac monitor use with respective orders

Pulmonary disorder (30.6%) followed by infectious disease (16.8%), gastroenterology (14.8%), and neurology (12.3%) related diseases were predominant admissions (Table1). These four systems constituted 74.5% of the admissions in this study cohort. The neurology group of patients was monitored without a physician order. On the other hand, the infectious disease group of patients had fewer monitoring and orders (Figure 1.B). Among these four systems, only gastroenterology order showed no statistical significance.

On average, all patients were admitted to the hospital for 2.8 days, and the median LOS was 1.7 days (IQR;1,3). One patient had the most prolonged stay (59 days), admitted for severe asthma with a protracted course. The length of hospital stays with either presence of monitors (cardiac monitor or pulse oximeter) or physician ordering for any of the two monitors was significant (Table 2). LOS was shorter in patients without monitors; the median difference for the cardiac monitor was 1 day (p = 0.002), and pulse oximeter was 0.5 day (p = 0.007). There was statistical significance in LOS among the “age” category (NICHD Pediatric Terminology[5], Figure 3, p = 0.046). The final model in multiple linear regression showed “cardiac monitor ordering” and “complex past medical history” had a significant effect on prolonging the length of stay (Table 2). Cardiac monitor order increased LOS by 22% (95% CI; 0.5%, 48%) whereas the presence of complex past medical history increased LOS by 25% (95% CI; 4%, 51%).

**Table 2.** Length of hospital stay (days) with various parameters

| Parameters | Mean(SD) | Median(IQR) | Parameters | Mean(SD) | Median(IQR) | p value* |
| --- | --- | --- | --- | --- | --- | --- |
| CM present | 3.2 (3.9) | 2 (1,3) | CM absent | 2.6 (5) | 1 (1,2) | 0.002 |
| PO present | 3.1 (3.9) | 2 (1,3) | PO absent | 2.6 (5) | 1.5 (1,2) | 0.007 |
| CM order (+) | 3.4 (4.2) | 2 (1,4) | CM order (-) | 2.6 (4.5) | 2 (1,2) | 0.0003 |
| PO order (+) | 3.4 (5.6) | 2 (1,3) | PO order (-) | 2.4 (2.5) | 2 (1,2.8) | 0.03 |

CM = cardiac monitor; PO = pulse oximeter; present (+), absent (-)
\*Wilcoxon Mann Whitney test

**Multiple linear regression (outcome = LOS, days)**
|  | Intercept | Cardiac monitor order | Complex past medical history |
| --- | --- | --- | --- |
| Coefficients | 3.467 | 0.2* | 0.226<sup>¶</sup> |
| Exponentiation | 32.06 | 1.22 | 1.25 |
| 95% confidence interval | 27.6, 37.3 | 1.005, 1.48 | 1.04, 1.51 |

\*p = 0.04 <sup>¶</sup>p = 0.02

**Figure 3.**
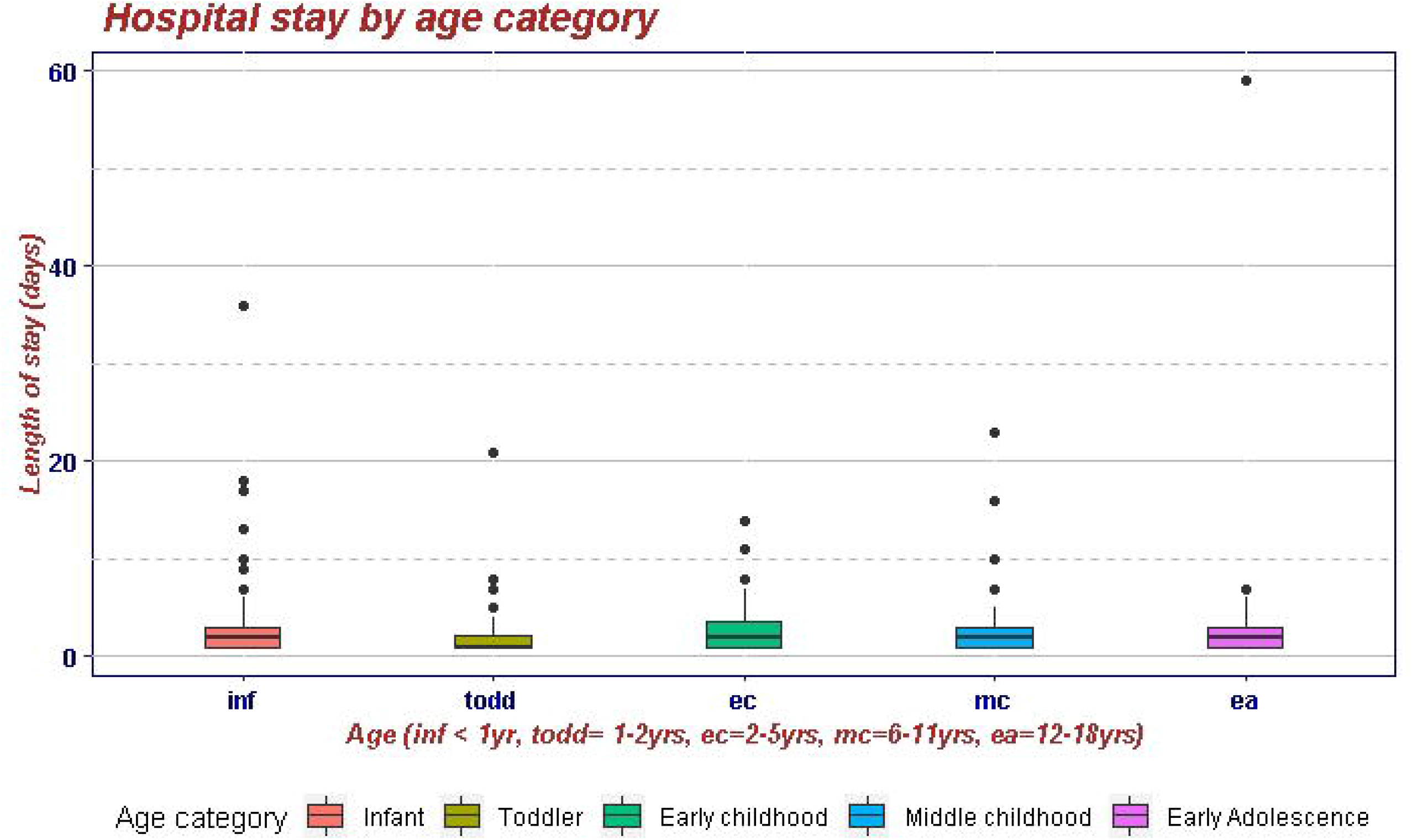
Effect of age on length of stay

## Discussion

We find that the variable practice of cardiac and pulse oximeter uses in pediatric patients in the general ward. The pulmonary group of patients was monitored more than the neurology group. More than half of pediatric admissions had both a cardiac monitor and pulse oximeter. A third of the patients did not have monitors. Our study showed the decision to monitor was associated with complexities in both past medical history and admission diagnosis. The patient’s age affects the decision to monitor. Hospital stay is reduced in patients without monitors. Multiple linear regression models showed complex medical conditions and cardiac monitor order prolonged hospital length of stay.

American Academy of Nursing (Choosing Wisely, April 19, 2018) recommended against continuous cardiac-respiratory or pulse oximetry monitoring to children and adolescents admitted to the hospital without appropriate indication. The nursing patient ratio differs according to the acuity of the ward patients. False alarms stress both clinicians and nurses, and fatigue is inevitable. Nurses can be distracted or desensitized due to alarms when multi-tasking. Monitoring guidelines and rationale use are available to develop consensus guidelines.[6,7] Guidelines for cardiac monitoring for pediatrics are limited[6], but efforts are made to reduce alarm fatigue.[8,9] Quality improvement projects address reducing alarm fatigue and nuisance in inpatient settings. [10-13]. More than half of the patients had monitors without orders (Figure1.A). Many hospitalists are developing consensus guidelines for monitoring based on weak evidence (email communication). Continuous physiologic monitoring must be tailored to patient type and location. Our study confirms that the pulmonary group of patients was most admissions, who will have monitoring. In pediatric wards, the decision is complex; stakeholders (patients, staff, ordering personnel), and system-level factors should be considered in local policy development.[14] A more holistic approach to understanding the patient’s condition, perspective of parents/caregivers, time of day, and setting expectations can reduce the overuse of monitors. Neurology patients had few orders to monitor in our study. This few-order raises uncertainty, e.g., whether patients with seizures or history of seizures need monitors? Hence, we suggest developing a standardized approach with the goal of patient safety and reduce harm.

Hypoxemia, either chronic or intermittent, is a concern. American Academy of Pediatrics (AAP) has provided guidelines for oxygen saturation by pulse oximeter above 90-92% in most healthy infants with bronchiolitis. Moreover, we extrapolate this acceptable saturation to common respiratory pediatric illnesses admitted to the hospital. [15,16]An evidence-based approach with practice standards and policy and continuing education can ensure appropriate use and reduce adverse events. More trials, such as reducing continuous pulse oximetry in bronchiolitis, will reduce the burden of overuse.[17] At the local level, there must be an ongoing dialog between caregivers, including parents, about the necessity of monitors, as the effectiveness of routine vital signs are unfounded.[18,19]

Most frequent admissions were predominantly respiratory, infections, neurology, and gastroenterology related disorders (Table 1). Continuous education on the utility of such technology and equipment for health care providers is warranted in this day and age.[20-23] With an emphasis on family-centered rounds, patient engagement can be counterproductive unless there is a clear understanding of using electronic monitoring.[24-26] Hendaus et al. showed that approximately 85% of caregivers feel safe monitoring with continuous pulse oximetry in children with bronchiolitis.[27] Similar kinds of literature related to bronchiolitis demonstrate the practice of monitoring without guidelines, and differing opinion influences threshold for admission[28,29], discharge[30], supplemental oxygen use[31-33], unnecessary test[34], and medication use. No study exists on the established benefit of electrocardiographic monitoring and telemetry use.[35,36] It is unclear if patients with non-life-threatening illnesses such as cellulitis, constipation with fecal impaction, bronchiolitis with no hypoxemia, and reduced oral intake benefit from cardiopulmonary monitoring.

Bivariate analysis showed complex medical history in the past or at admission influence the decision to order the monitor. Complex past medical history increased LOS by 25% in multiple linear regression. This increase in LOS can be explained by the fact that some children are technology dependent, trainee experience, and parent expectation of a hospitalized sick child. In a retrospective study by Gold et al., 14.9% of children with medical complexity stayed for more than 10day.[37] Judicious use of monitoring, considering what is monitored at home versus adding more in the hospital admission, can make a difference. Efforts such as “Compass care,” a care-coordination without compromising the quality of care for children with medical complexity by Children’s Hospital Association partners, had reduced LOS from 4.76 days per patient per month in the year to 1.26 days. Age can be confounding in this decision-making process, as noted in figure 2, but we did not look at the individual patient level for medical complexity.

In this study, we showed that the cardiac monitor order was associated with a 22% increased length of stay. Interventions by hospitalists to improve appropriate use can reduce LOS and cost.[38]

Our study result is strengthened by the authors physically checking the orders and presence of monitors on the patients in real-time. We did not check with the nurses if they intended to follow the orders or use their judgment. Patients would have received monitoring orders in the latter part of the day when clinical condition changed, or physician remembered to order by default. Our study did not account for the cost involved, but it is a medical waste if no indication for the monitor. We did not analyze why monitors were used on patients other than non-cardiorespiratory or with airway problems.

The inpatient monitoring needs standardization. Guidelines to monitor, for example, can be stratified by organ systems, patient groups, and patient location within the hospital. Without clear guidelines for either cardiac or pulse oximeter monitoring, overdiagnosis (e.g., relative hypoxemia, bradycardia), over-testing, and over-use of technology are cumbersome. The monitoring of patients must be cost-effective and safe.

## Data Availability

Data not available.

